# Declined functional connectivity of white matter during rest and working memory tasks associates with cognitive impairments in schizophrenia

**DOI:** 10.1101/2020.05.16.20091397

**Authors:** Yurui Gao, Muwei Li, Anna S. Huang, Adam W. Anderson, Zhaohua Ding, Stephan H. Heckers, Neil D. Woodward, John C. Gore

**Author notes:** Equal contributions (Neil D. Woodward, John C. Gore).

## Abstract

**BACKGROUND:** 

Schizophrenia, characterized by cognitive impairments, arises from a disturbance of brain network. Pathological changes in white matter (WM) have been indicated as playing a role in disturbing neural connectivity in schizophrenia. However, deficits of functional connectivity (FC) in individual WM bundles in schizophrenia have never been explored; neither have cognitive correlates with those deficits.

**METHODS:** 

Resting-state and spatial working memory task fMRI images were acquired on 67 healthy subjects and 84 patients with schizophrenia. The correlations in blood-oxygenation-level-dependent (BOLD) signals between 46 WM and 82 gray matter regions were quantified, analyzed and compared between groups under three scenarios (i.e., resting state, retention period and entire time of a spatial working memory task). Associations of FC in WM with cognitive assessment scores were evaluated for three scenarios.

**RESULTS:** FC deficits were significant (*p*<.05) in external capsule, cingulum, uncinate fasciculus, genu and body of corpus callosum under all three scenarios. Deficits were also present in the anterior limb of the internal capsule and cerebral peduncle in task scenario. Decreased FCs in specific WM bundles associated significantly (*p*<.05) with cognitive impairments in working memory, processing speed and/or cognitive control.

**CONCLUSIONS:** Decreases in FC are evident in several WM bundles in patients with schizophrenia and are significantly associated with cognitive impairments during both rest and working memory tasks. Furthermore, working memory tasks expose FC deficits in more WM bundles and more cognitive associates in schizophrenia than resting state does.

## INTRODUCTION

Schizophrenia, a disorder characterized by cognitive impairments (1-3), has been long hypothesized to arise from a ‘disconnection’ of neural networks (4, 5). This hypothesis is supported by neuroimaging studies showing the alteration of functional connectivity (FC) between gray matter (GM) regions and its link to cognitive impairment in schizophrenia, common within fronto-temporal (6), fronto-parietal (7, 8), fronto-hippocampal (9) and thalamo-cortical (10, 11) circuits.

FC between GM regions is usually examined using functional magnetic resonance imaging (fMRI). Specifically, temporal correlation between blood-oxygen-level-dependent (BOLD) signals from a pair of GM regions is interpreted as an indicator of FC between them. By contrast, FC in white matter (WM), another fundamental component to form a neural network, has been ignored for decades (12). That is because BOLD signals in WM are weaker due to less blood flow or volume (13) and consequently are usually excluded from image analyses. However, emerging fMRI evidence has demonstrated that BOLD effects in WM are robustly detectable (14-18). Our recent work has shown that BOLD signals from WM bundles correlate with those from specific GM regions to which they connect (19, 20). Based on this correlation, we derived a WM-centered FC analysis approach and revealed that the FC at specific WM bundles declined and associated with cognitive scores in Alzheimer’s disease during resting state (21, 22).

To our knowledge, this WM-centered FC analysis approach has not been applied to research of schizophrenia and may yield novel insights into brain network dysfunction and possible mechanisms of cognitive impairment in schizophrenia. Moreover, anatomical changes of WM found in schizophrenia, such as alterations of myelin sheath lamellae (23, 24) and decrease in WM integrity (25-28) or volume (29-32), are likely to provide a pathophysiological explain for possible FC changes in WM. With these motives, we hypothesize that FC in WM, whether during rest or task, may alter in schizophrenia and associate with cognitive functioning.

Furthermore, our prior work also showed that the FC between WM and GM can be modulated by functional loading such as visual activation in healthy subjects (19), which raises a question in general: whether the detected FC alteration in WM in disease can be changed by functional loading (e.g., induced by a task). However, the question has not been answered in any of prior studies.

Accordingly, in this study we extend our previous analyses (19, 21, 22) to a large cohort of patients with schizophrenia and healthy controls on both spontaneous signal during a resting state and activated signal during a spatial working memory task in order to: 1) quantitatively characterize the alterations of FC in WM affected by schizophrenia; 2) explore associations between FC in WM and cognitive functioning across the cohort; 3) further determine whether the FC alterations at WM and cognitive associates detected in schizophrenia are changed by cognitive functional loading.

## MEMTHODS AND MATERIALS

### Participants

Sixty-seven healthy controls (CON) and 84 patients with schizophrenia spectrum disorders (SCZ) (Table 1) were recruited at Vanderbilt Psychiatric Hospital. The SCZ patients included individuals diagnosed with schizophrenia (n=51), schizoaffective disorder (n=10) and schizophreniform disorder (n=23). The Structured Clinical Interview for DSM-IV Disorders (33) was administered to confirm diagnoses in patients and rule out current or past psychiatric illnesses in healthy participants. Patients were further assessed with the Positive and Negative Syndrome Scale (PANSS) (34) to quantify severity of clinical symptoms. Study procedures and exclusion criteria are described in detail in the Supplement. This study was approved by the Vanderbilt University Institutional Review Board and all participants provided written informed consent.

**Table 1.**
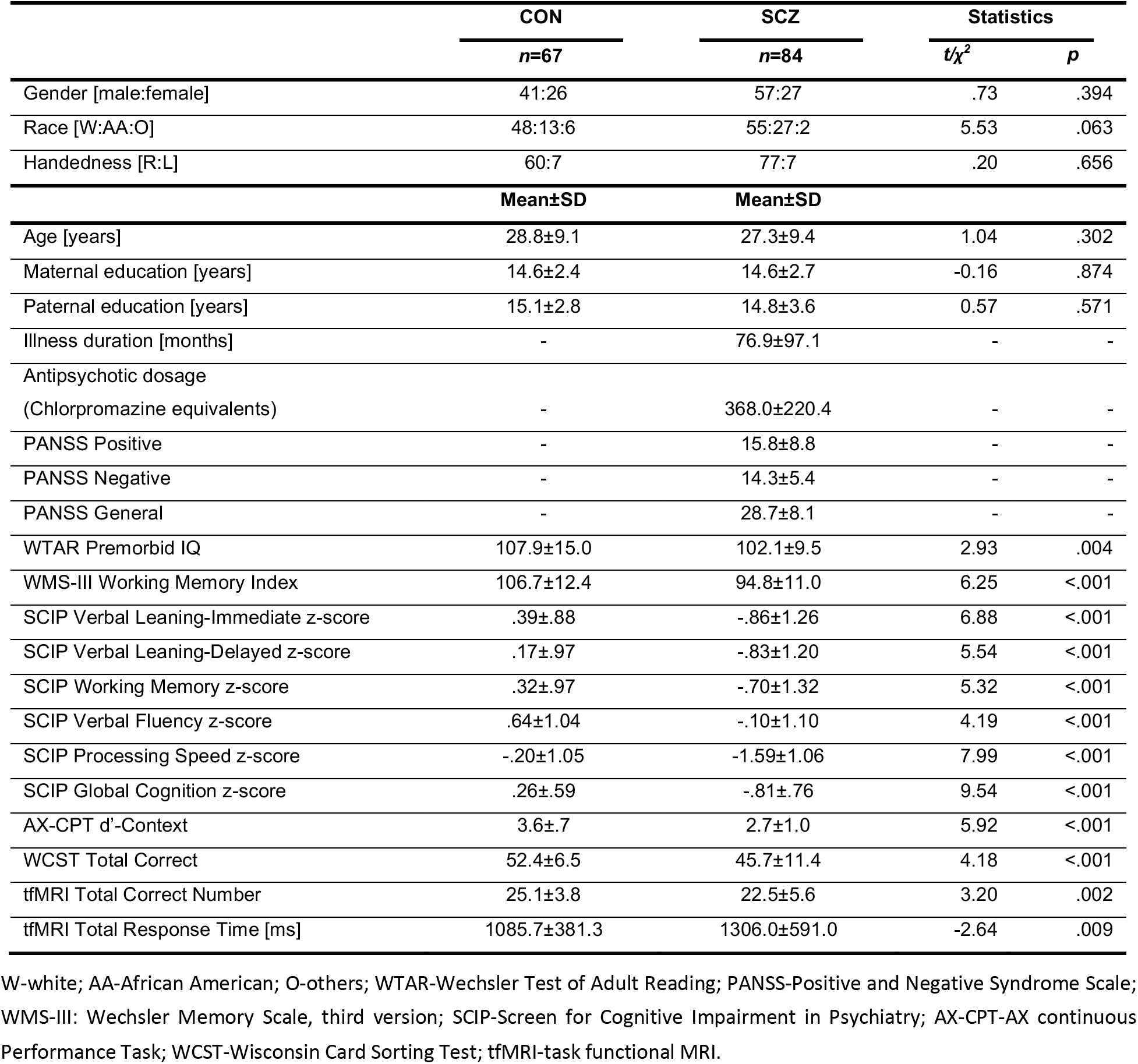
Demographics, clinical and cognitive characteristics

### Cognitive Assessments

Each participant was administered the same cognitive tests described as follows. The Wechsler Test of Adult Reading (WTAR; (35)), a single word-reading test, was performed to estimate premorbid intellect. The spatial span and letter-number sequencing subtests from Wechsler Memory Scale-3rd edition (WMS-III; (36)) were completed and together yielded the working memory index. The administered Screen for Cognitive Impairment in Psychiatry (SCIP; (37)) included a word list learning test of verbal memory, a version of the auditory consonant trigrams test of working memory, phonemic verbal fluency and a coding test of processing speed. SCIP subtests raw scores were converted to z-scores and averaged to create a global cognition z-score. The AX Continuous Performance Task (AX-CPT; (38)) was administered and the d-prime (d’), referred to as d’-context, was computed from the AX hits and BX false alarm (39). In addition, the Wisconsin Card Sorting Test (WCST; (40)), a test of executive function, was scored. All the cognitive assessment scores acquired in this study are listed in Table 1.

### Spatial Working Memory Task

Each working-memory task fMRI scan comprised 5 spatial working memory trials and 3 non-memory-related trials (Figure 1A). Subjects were instructed to remember positions of three spatial locations in the memory trial and performed a sensorimotor task devoid of memory requirements in the non-memory-related trial. Each trial started with a 4-second fixation, followed by three dots appearing sequentially within the next 3.5 seconds. After a 16-second retention interval, a probe stimulus of 1 second appeared, and the subjects responded to a probe location with a button press indicating whether this stimulus was at one of the memorized locations. At the end of each trial, there was an inter-trial interval of 13.5 seconds that included the response to the stimulus (Figure 1B). The non-memory-related trial was identical except subjects were instructed not to remember anything but simply to press both buttons when the probe appeared. Different colored dots were used to cue subjects to working memory trial (red dots) or non-memory trial (grey dots). The BOLD signal for healthy controls from Brodmann area 47 in each hemisphere during one spatial working memory trial is shown in Figure 1C. More details were described in the previous study (41). The total correct number and total response time were recorded to measure performance of this spatial working memory task, as listed in Table 1.

**Figure 1.**
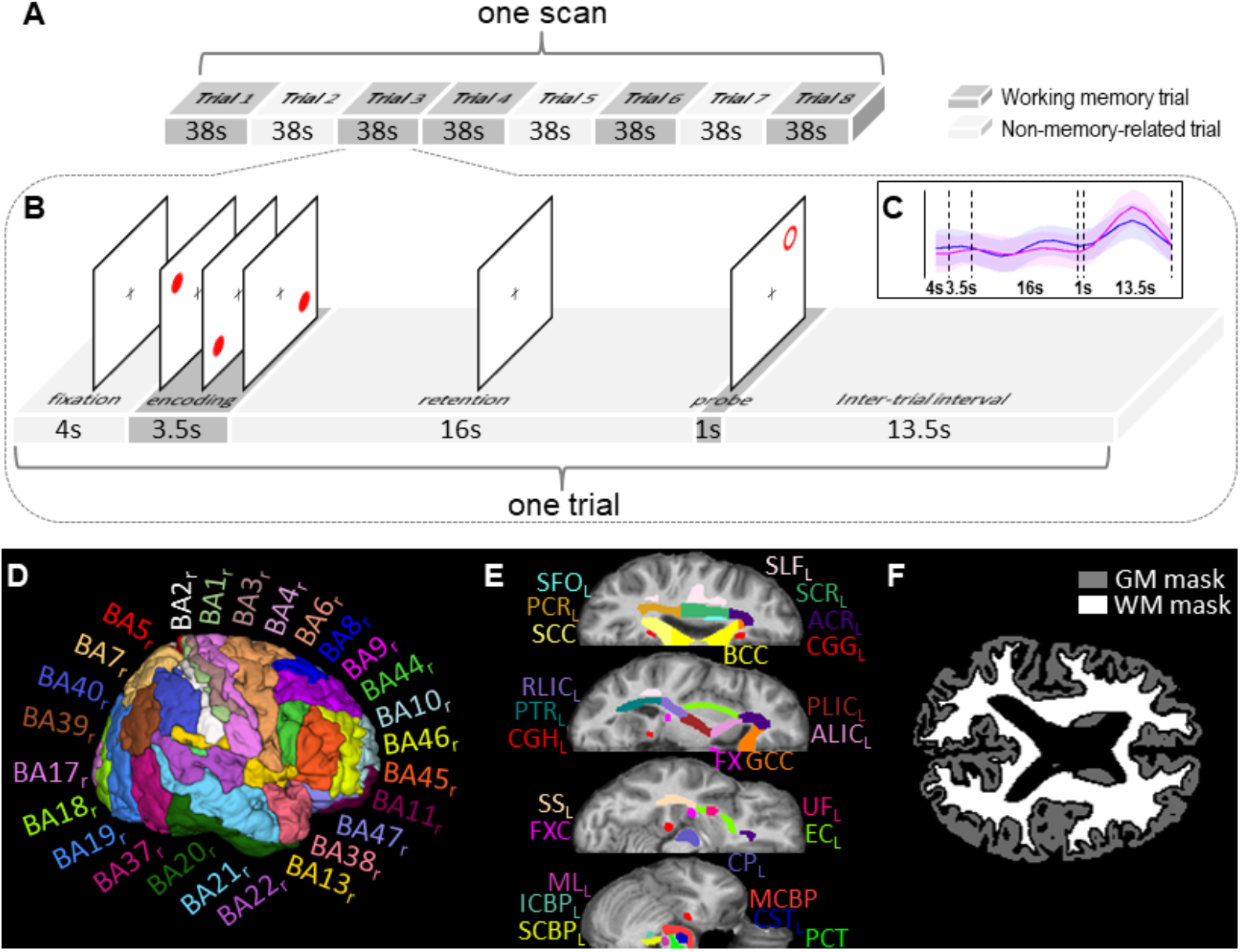
Schematic diagram of spatial working memory task events, atlases of white matter (WM)/ gray matter (GM) ROIs, and tissue masks. **(A)** One task fMRI scan comprised eight trials, of which five were working memory trials (dark gray blocks) and the remaining three were non-memory-related trials (light gray blocks). **(B)** Each memory trial lasted 38 seconds, including 4 seconds of fixation, 3.5 seconds of encoding, 16 seconds of retention, 1 second of stimulus and 13.5 seconds of inter-trial interval. The non-memory trial had the same sequence of events, except subjects were instructed not to remember the target locations. **(C)** Averaged BOLD signal of all healthy controls on Brodmann area 47 during one working memory trial. **(D)** GM parcellation atlas and **(E)** WM parcellation atlas in MNI space were used to initially define WM and GM ROIs. See Table 2 for lists of these ROIs. **(F)** GM and WM whole-brain tissue masks of one healthy subject used to further constrain GM and WM ROIs to avoid signal contamination between any GM ROI and its adjacent WM ROIs.

### MRI Image Acquisition and Preprocessing

One resting state fMRI scan (sequence=echo-planar imaging, TR/TE=2s/35ms, resolution=3×3×3mm^3^, matrix= 80×80×38, dynamics per scan=300, eyes closed), six working-memory task fMRI scans (same parameters except dynamics per scan=152) and one T1-weighted scan (sequence=turbo field echo, TR/TE=8ms/3.7ms, resolution= 1×1×1mm^3^, matrix=256×256×170) were acquired for each subject using one of two identical 3T MRI scanners (Philips Healthcare Inc., Best, Netherlands) with 32-channel head coils at Vanderbilt University Institute of Imaging Science.

Image preprocessing is described in detail in the Supplement. Briefly, preprocessing of fMRI images included correcting slice timing and head motion, regressing out 24 motion parameters and mean cerebrospinal fluid (CSF) signal, temporal filtering (passband=.01-.1Hz), co-registering to the Montreal Neurological Institute (MNI) space, detrending, and voxel-wise normalization of the time-courses into zero mean and unit variance. In order to avoid signal contamination between WM and GM in preprocessing, we did not spatially smooth fMRI data. Preprocessing of T1-weighted images included segmenting WM, GM, and CSF and co-registering the resultant tissue probability maps to the MNI space.

### Analyses of Functional Connectivity in White Matter

All the analyses were based on functional correlation matrix (FCM) - a matrix of correlations between WM and GM regions of interests (ROIs). The WM and GM ROIs were initially defined by the Eve atlas (42) (46 WM bundles in both cerebrum and cerebellum, Figure 1E and Table 2) and PickAtlas (43) (82 Brodmann areas in cerebral cortex, Figure 1D and Table 2), respectively. Those WM or GM ROIs were further constrained within WM or GM whole-brain masks generated by thresholding the WM or GM tissue probability maps at 0.8 (Figure 1F) in order to avoid signal contamination between WM and GM. The preprocessed time-courses were averaged over each ROI. Each two averaged time-courses from one pair of WM and GM ROIs were then linearly correlated, excluding any time points with large head motions (i.e., frame-wise displacement (44) >.5). The resulting 46×82 correlation coefficients comprised an FCM of WM-GM pairs. The possible influences of gender, race, age, maternal and paternal years of education were regressed out from FCM using a generalized linear model.

**Table 2.** List of white matter and gray matter ROIs (BA: Brodmann area).

| **White Matter (WM) ROIs** | **Gray Matter (GM) ROIs** |
| --- | --- |
| CST: Corticospinal Tract | BA1: Primary Somatosensory Cortex 1 |
| ML: Medial Lemniscus | BA2: Primary Somatosensory Cortex 2 |
| ICBP: Inferior Cerebellar Peduncle | BA3: Primary Somatosensory Cortex 3 |
| SCBP: Superior Cerebellar Peduncle | BA4: Primary Motor Cortex |
| CP: Cerebral Peduncle | BA5: Somatosensory Association Cortex |
| ALIC: Anterior Limb of Internal Capsule | BA6: Premotor and Supplementary Motor |
| PLIC: Posterior Limb of Internal Capsule | BA7: Visuo-Motor Coordination |
| RLIC: Retrolenticular Limb of Internal Capsule | BA8: Frontal Eye Fields |
| ACR: Anterior Corona Radiata | BA9: Dorsolateral Prefrontal Cortex |
| SCR: Superior Corona Radiata | BA10: Anterior Prefrontal Cortex |
| PCR: Posterior Corona Radiata | BA11: Orbitofrontal Area |
| PTR: Posterior Thalamic Radiation (include Optic Radiation) | BA13: Insular Cortex |
| SS: Sagittal Stratum (include inferior longitudinal fasciculus and fronto-occipital fasciculus) | BA17: Primary Visual Cortex (V1) |
| EC: External Capsule | BA18: Secondary Visual Cortex (V2) |
| CGG: Cingulum (Cingulate Gyrus) | BA19: Associative Visual Cortex (V3-5) |
| CGH: Cingulum (Hippocampus) | BA20: Inferior Temporal Gyrus |
| FXC: Fornix (Cres) | BA21: Middle Temporal Gyrus |
| SLF: Superior Longitudinal Fasciculus | BA22: Superior Temporal Gyrus |
| SFO: Superior Fronto-Occipital Fasciculus | BA23: Ventral Posterior Cingulate Cortex |
| UF: Uncinate Fasciculus | BA24: Ventral Anterior Cingulate Cortex |
| MCBP: Middle Cerebellar Peduncle | BA25: Subgenual Area |
| PCT: Pontine Crossing Tract | BA26: Ectosplenial Portion of Retrosplenial Region |
| GCC: Genu of Corpus Callosum | BA27: Piriform Cortex |
| BCC: Body of Corpus Callosum | BA28: Ventral Entorhinal Cortex |
| SCC: Splenium of Corpus Callosum | BA29: Retrosplenial Cingulate Cortex |
| FX: Fornix | BA30: Part of Cingulate Cortex |
|  | BA32: Dorsal Anterior Cingulate Cortex |
|  | BA34: Dorsal Entorhinal Cortex |
|  | BA35: Perirhinal Cortex |
|  | BA36: Ectorhinal Area |
|  | BA37: Occipitotemporal Area (part of fusiform gyrus and inferior temporal gyrus) |
|  | BA38: Temporopolar Area |
|  | BA39: Angular Gyrus |
|  | BA40: Supramarginal Gyrus |
|  | BA41: Auditory Cortex 1 |
|  | BA42: Auditory Cortex 2 |
|  | BA43: Primary Gustatory Cortex |
|  | BA44: Pars Opercularis |
|  | BA45: Pars Triangularis |
|  | BA46: Dorsolateral Prefrontal Cortex |
|  | BA47: Pars Orbitalis |

To analyze FC alterations in schizophrenia relative to normal, we averaged the FCMs across subjects within each group, denoted as mFCM, and then subtracted mFCM of SCZ group from mFCM of CON group by element-wise subtraction. Unpaired *t*-test was conducted for each FCM element to determine the significance of this inter-group difference in FC. The resulting 46×82 *p*-values were corrected for multiple comparisons using a false discovery rate (FDR) (45), denoted as *p*_FDR_. The effect size of difference (46) for each FCM element between groups was also calculated. To estimate the overall FC of each WM bundle, the 82 FCM elements subject to each WM ROI were averaged. The overall FCs of each WM-tract across all subjects were then group-averaged and the group data were compared using unpaired *t*-tests.

### Association with Cognitive Scores

The association between each single FCM element, i.e., FC of one WM-GM pair, and each cognitive score was evaluated by calculating the Pearson’s correlation coefficient, r, between them across all subjects. The 46×82 resulting *p*-values for each score were corrected using FDR. In this way, given one score, each WM bundle had 82 correlation coefficients. Among those coefficients, the one with maximum amplitude was selected to represent the association between the FC of the WM bundle and the score. To make the result even more statistically rigorous, given one score and one WM bundle, if there is only one coefficient meets with *p*_FDR_ <.05 out of 82 coefficients, suggesting that it is hard to decide whether this significance appeared by chance or not, then this WM bundle was determined not to associate with the score.

### Three Scenarios

To examine the influence of functional loading on detection of FC alterations in WM and their cognitive associates, three FCMs were calculated using the time-courses acquired in three scenarios: ‘resting state’, ‘working memory I’, and ‘working memory II’. In the resting state scenario, each time-course included all eligible images from resting state fMRI scan, yielding FCM_RS_. In the working memory I scenario, each time-course included only the images acquired during the retention periods of all 5 memory trials of all 6 task scans, yielding FCM_WMI_. In the working memory II scenario, each time-course included the images acquired throughout the entire time of all 5 memory trials of all 6 task scans, yielding FCM_WMII_. For each scenario, the analyses described above were performed separated.

## RESULTS

### Participant Characteristics

Table 1 summarizes the demographic, clinical, and cognitive characteristics of all 151 participants from the CON group (n=67) and SCZ group (n=84). Between the two groups, no significant differences in age (*p*=.302), gender (*p*=.394), race (*p*=.063), handedness (*p*=.656), maternal education (*p*=.874) and paternal education (*p*=.571) were observed. As anticipated, all the cognitive and fMRI task scores were significantly different between the two groups (*p*<.01).

### Alterations in White Matter Functional Connectivity

Figure 2 shows the mFCM for the CON and SCZ groups in the three scenarios: resting state, working memory I and working memory II. The general patterns of mFCM across the three scenarios appear similar, but the two working memory scenarios exhibit stronger FCs in some WM bundles, e.g., anterior limb of internal capsule (ALIC), particularly clear in the ALIC-BA6 pair. Moreover, compared with mFCM of the CON group in each scenario, the corresponding mFCM of the SCZ group appears to have a generally weaker FC pattern.

The element-wise differences of mFCM between the CON and SCZ groups in the three scenarios are shown in Figure 3A-C, where only the elements with significant differences (*p*_FDR_<.05) are presented with non-zero values. Clearly, the SCZ group has reduced FCs at several WM-GM ROI pairs but also increased FCs at a few WM-GM pairs relative to the CON group in each scenario. The effect sizes of the group differences for the elements with *p*_FDR_ <.05 were all higher than 0.45 in all three scenarios, as shown in Supplemental Figure S1, indicating that the differences are not trivial. Moreover, among the three scenarios, working memory II reveals FC reductions at more WM-GM ROI pairs.

**Figure 2.**
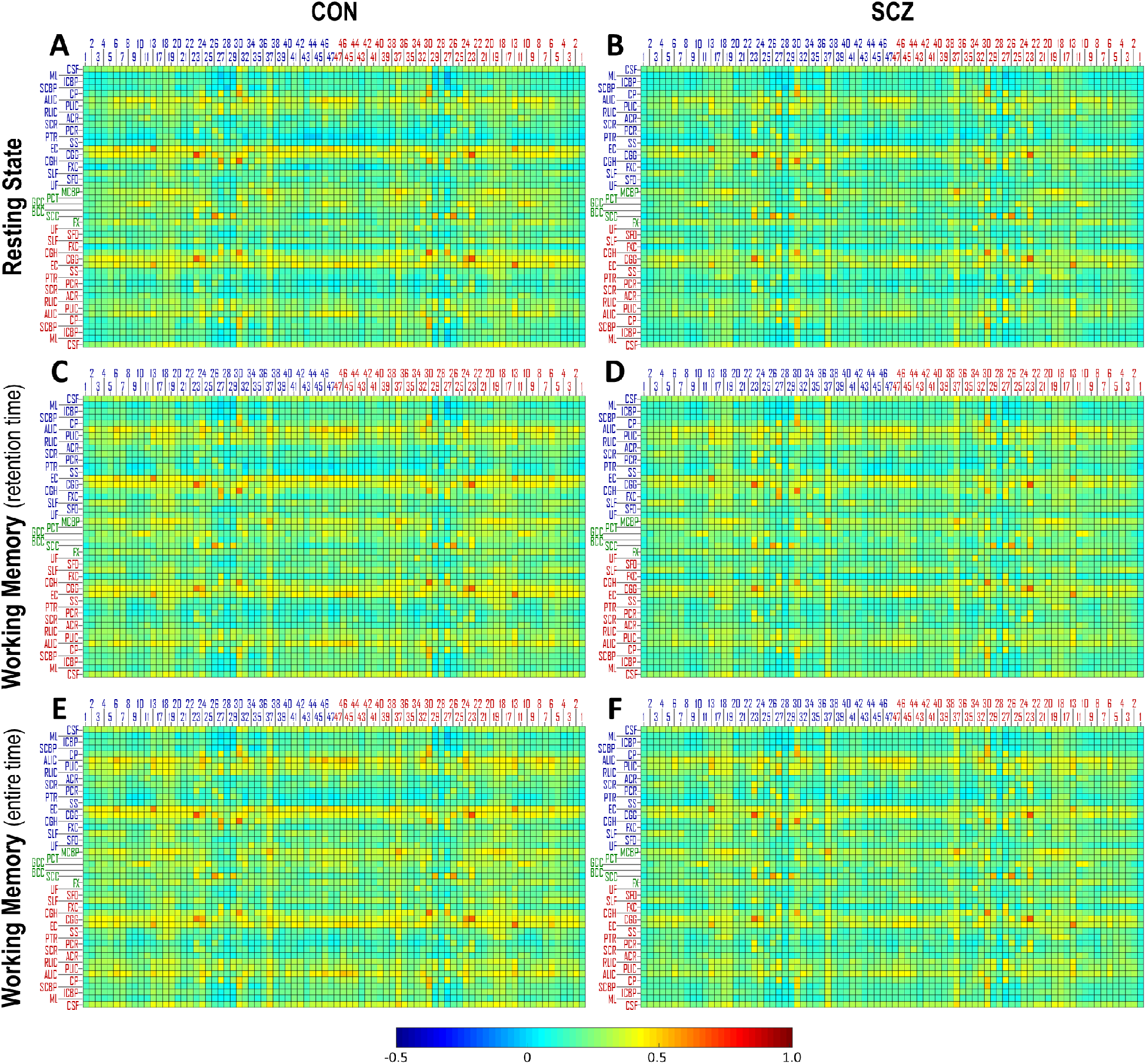
Group mean of functional correlation matrix (mFCM) for CON group and SCZ group in three scenarios: **(A, B)** resting state, **(C, D)** working memory **I** and **(E, F)** working memory **II**. The blue/red index labeling each column of mFCM indicates the number of Brodmann area in left/right hemisphere. The blue/green/red abbreviation labeling each row of mFCM indicates the WM bundle in left/middle/right portion of brain. See Table 2 for lists of these ROIs.

The group comparison of the FC in each of the 46 WM bundles under each of the three scenarios is presented in Figure 3D and Supplemental Table S1. Under resting state, the FC declines in SCZ relative to CON are highly significant in bilateral external capsule (EC; both *p* <.001), bilateral uncinate fasciculus (UF; *p* =.006 and *p* =.003, respectively), genu of corpus callosum (GCC; *p* =.008) and body of corpus callosum (BCC; *p* =.006) and significant at bilateral cingulum near cingulate gyrus (CGG; *p* =.017 and *p* =.013, respectively). Meanwhile, SCZ data show a significant increase in left posterior thalamic radiation (PTR; *p* =.031). Under working memory I scenario, highly significant reductions in SCZ relative to CON were found at left EC (*p* =.006) and right fornix cres (FXC; *p* =.009), and significant reductions were found in bilateral CGG (*p* =.013 and *p* =.020, respectively), left UF (*p* =.017), GCC (*p* =.046), BCC (*p* =.014), right cingulum near hippocampus (CGH; *p* =.013) and right EC (*p* =.017). Right UF (*p* =.069) exhibited a trend to a significant reduction. By contrast, right posterior corona radiata (PCR) showed a significant increase (*p* =.026) in SCZ relative to the CON. Under working memory II scenario, we found highly significant decreases in SCZ relative to CON in bilateral EC (*p* <.001 and *p* =.006, respectively), bilateral CGG (both *p* =.007), left UF (*p* =.007), right FXC (*p* =.006) and right CGH (*p* =.005). Significant decreases were found in GCC (*p* =.017), BCC (*p* =.014), right UF (*p* =.011), right ALIC (*p* =.045) and right cerebral peduncle (CP; *p* =.029).

**Figure 3.**
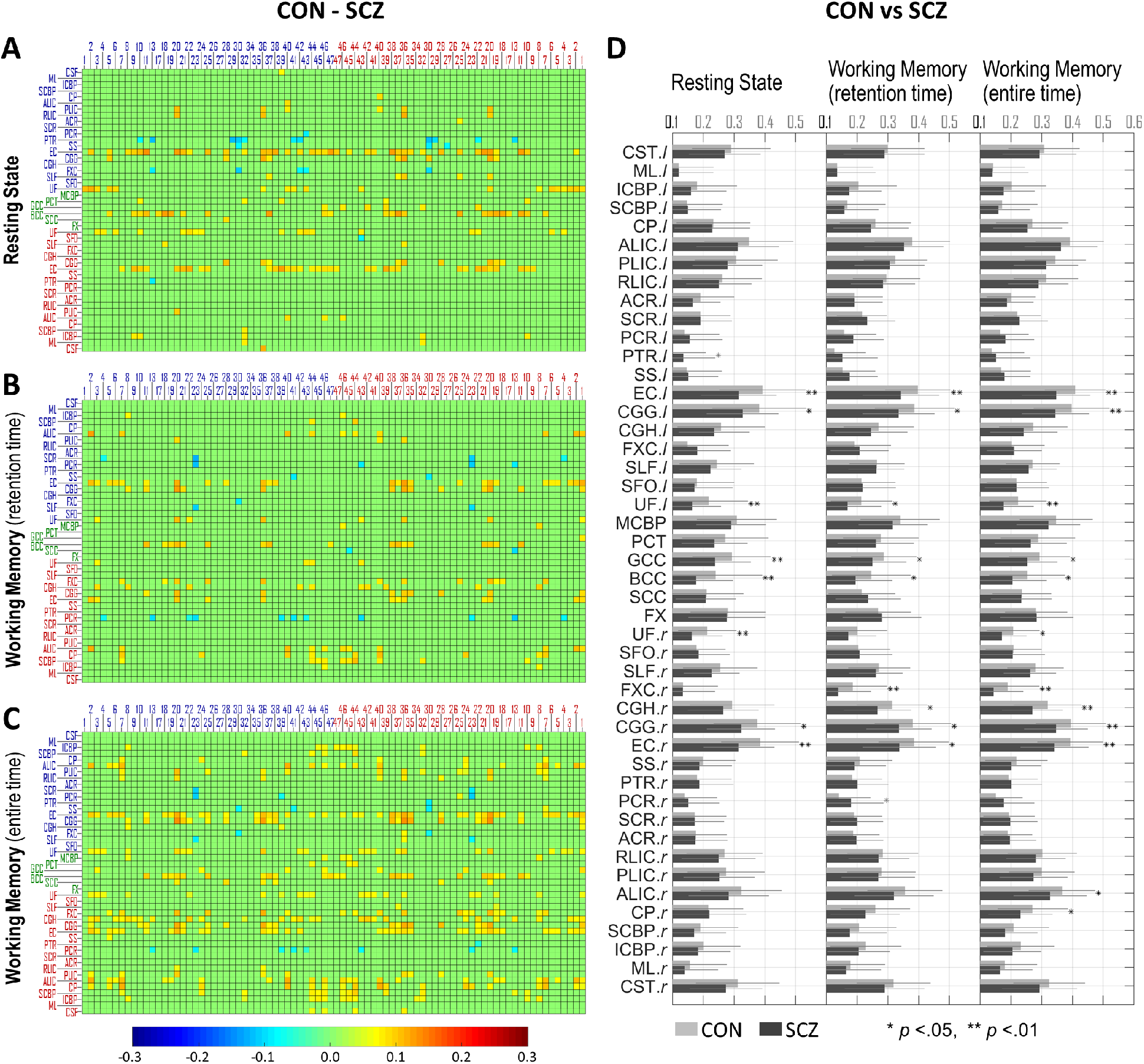
Comparisons of FCM and FC in WM bundle between CON group and SCZ group in three scenarios. **(A-C)** Difference of subtracting mFCM of SCZ from mFCM of CON in three scenarios: **(A)** resting state, **(B)** working memory I and **(C)** working memory II. The differences of FCM elements with *p*_FDR_>.05 were set to zero. The blue/red indices labeling each column indicate number of Brodmann areas in left/right hemispheres. The blue/green/red abbreviations labeling each row indicate WM bundles in left/middle/right of brain. See Table 2 for the lists of these ROIs. **(D)** Group means of FC in WM bundle for CON group (light gray bar) and SCZ group (dark gray bar) in each of three scenarios. * indicates *p* <.05 and ** indicates *p* <.01. See Supplement Table S1 for group mean, standard deviation and *p*-values of each WM bundle as a quantitative reference.

### Associations between White Matter Functional Connectivity and Cognitive Scores

The significant associations (thresholding criteria: (*p*_FDR_ <.05 and |r| >.3) or (*p*_FDR_ <.01 and | bundles and cognitive scores (including fMRI task scores) are depicted in Figure 4.C Working Memory II (entire time)

**Figure 4.**
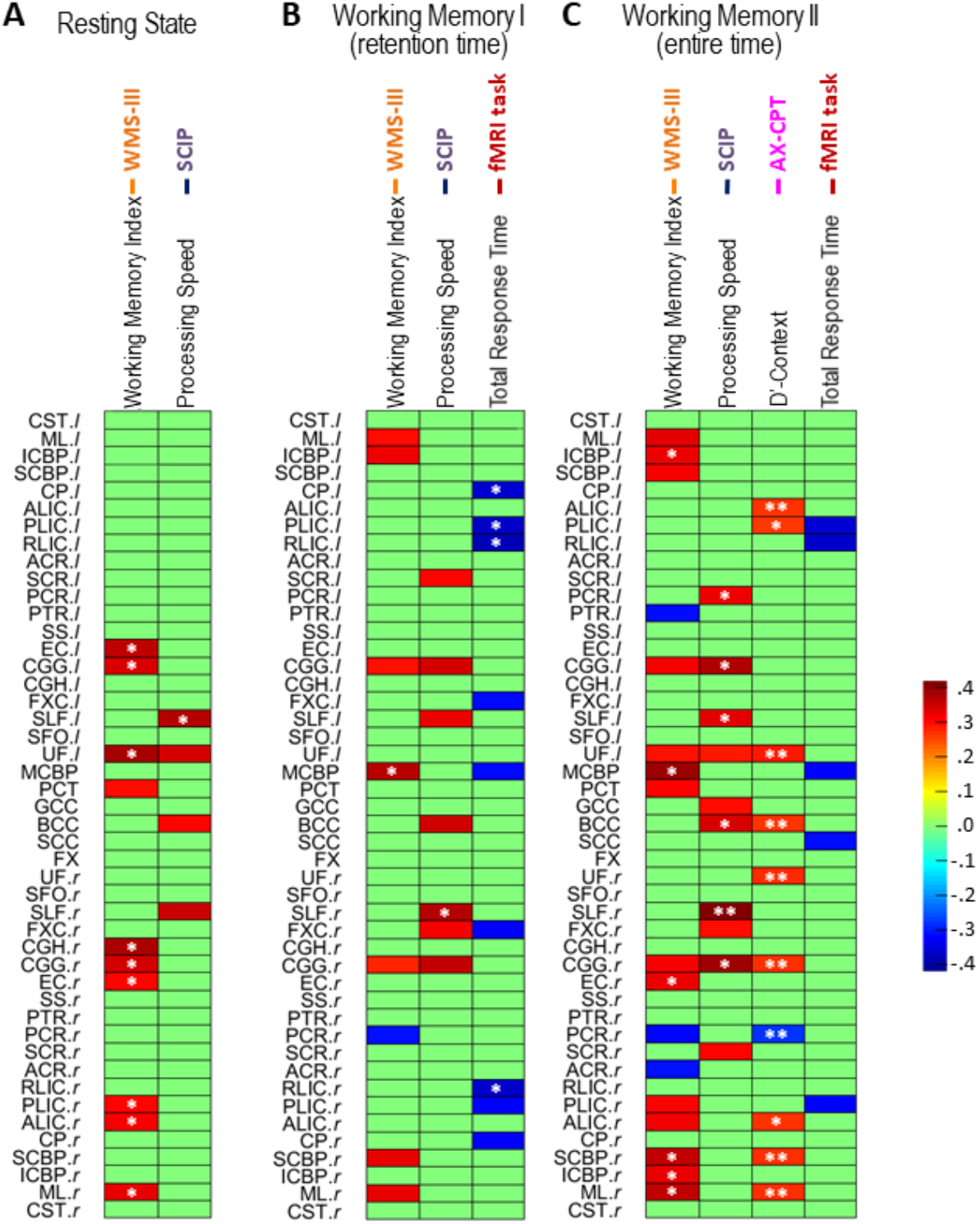
Correlations between FCs and cognitive scores in three scenarios: **(A)** resting state, **(B)** working memory I and **(C)** working memory II. Each correlation corresponds to one WM bundle and one score, and the correlation value is the one with maximum amplitude among all between the score and all 82 FCs of this WM bundle with 82 GM ROIs. All the non-zero values are with p<.05 (* indicates p<.01, ** indicates p<.001).

In resting state (Figure 4A), the FC in some WM bundles positively correlated with WMS-III working memory index or SCIP processing speed z-score. In particular, the highly significant correlations (*p*_FDR_ <.01 and r >.3) were found with WMS-III working memory index in bilateral EC, bilateral CGG, left UF, right CGH, right posterior limb of internal capsule (PLIC), right ALIC and right medial lemniscus (ML), with SCIP processing speed in left superior longitudinal fasciculus (SLF). There were no significant correlations (*p*_FDR_ <.05) between FC and other cognitive scores.

In working memory I scenario (Figure 4B), significant associations were found with WMS-III working memory index, SCIP processing speed z-score or total response time of fMRI task. Specifically, WMS-III working memory index positively correlated (*p*_FDR_ <.05 and r >.3) with FC in bilateral ML, left interior cerebellar peduncle (ICBP), bilateral CGG, middle cerebellar peduncle (MCBP) and right superior cerebellar peduncle (SCBP). SCIP processing speed positively correlated (*p*_FDR_ <.05 and r >.3) with FC at left superior corona radiata (SCR), bilateral CGG, bilateral SLF, BCC, and right FXC. Total response time of fMRI task negatively correlated (*p*_FDR_ <.05 and r < -.3) with FC at bilateral CP, bilateral PLIC, bilateral retrolenticular limb of internal capsule (RLIC), bilateral FXC and MCBP.

In working memory II scenario (Figure 4C), significant correlations were found with WMS-III working memory index, SCIP processing speed z-score, AX-CPT accuracy d’ or total response time of fMRI task. In particular, WMS-III working memory index positively correlated (*p*_FDR_ <.05 and r >.3) with FC at bilateral ML, bilateral ICBP, bilateral SCBP, bilateral CGG, left UF, MCBP, pontine crossing tract (PCT), right EC, right PLIC, and right ALIC. SCIP processing speed positively correlated (*p*_FDR_ <.05 and r >.3) with FC at left PCR, bilateral CGG, bilateral SLF, left UF, GCC, BCC, right FXC and right SCR. AX-CPT d’ context highly positively correlated (*p*_FDR_ <.01 and r >.2) with FC at bilateral ALIC, left PLIC, bilateral UF, BCC, right CGG, right SCBP and right ML. Total response time of fMRI task negatively correlated (*p*_FDR_ <.05 and r < -.3) with FC at bilateral PLIC, left RLIC, MCBP and SCC.

## DISCUSSION

We revealed that the FC declined at several WM bundles in patients with schizophrenia relative to healthy controls and that the declined FC at several WM bundles significantly associated with impaired cognition involving working memory, processing speed and cognitive control. Furthermore, comparison of the results across three scenarios confirmed that the FC alterations at WM bundles and their cognitive associates detected in schizophrenia varied depending on the functional loading onto brain. Together, these findings support a role for the WM FC deficits in cognitive impairments and suggest that FC at WM may serve as an additional biomarker of network change in schizophrenia. This study provides a first look at FC alterations at WM bundles and their associations with cognitive functioning in a large group of patients with schizophrenia and heathy controls.

The derivation of FC values for WM-GM pairs follow the same methodology as that used to infer FC between cortical regions and identify brain networks, but the interpretation of FCMs remains not fully clear (for a recent review see Gore et al. (15)). There are potential concerns of the influence of residual GM partial volumes in WM voxels, though these are likely insignificant using the methods adopted in this study, which included applying a highly conservative mask to constraint ROIs, using a deep WM atlas and avoiding spatial smoothing. From other studies, BOLD signals in WM are clearly associated with neural activity changes (15, 16) but it is not clear whether those changes are intrinsic to WM or arise from neighboring GM and potentially reflect vascular drainage effects from the cortex. Millan *et al*. found that the deep venous system draining deoxygenated blood in deep WM is separate from the superficial venous system draining blood in GM and superficial WM (47), suggesting that BOLD signal in deep WM is unlikely to be affected by vascular changes in GM.

Whether loading brain network with the spatial working memory task or not, the FC declines in SCZ group were consistently detected at EC, CGG, UF, GCC and BCC (Figure 3D), whose fractional anisotropy reductions were reported in prior diffusion MRI studies on schizophrenia (25, 27, 48, 49). This indicates that these bundles fail to function normally in patient’s brain network even at rest and this failure is likely due to the intrinsic structural degradation occurring in these bundles. By contrast, FC declines in FXC and CGH (Figure 3D) were detected under working memory scenarios but not at rest. Previous lesion studies have demonstrated that fornix and cingulum participate in working memory functioning (50, 51). These findings hint that loading brain network with a proper function may reveal FC deficits in the WM bundles that are engaged in this function. The FC declines in ALIC and CP, found in working memory II scenario only (Figure 3D), may be related to the onset of the stimulus response (Supplement Figure S2). Comparing memory II scenario with I (Figure 3C,D), the ALIC-paired GM ROIs with a significantly decreased FC only in memory II scenario included BA5 (superior parietal lobe), BA18/19 (visual cortex V2-V5), BA24 (anterior cingulate cortex) and BA37 (fusiform gyrus), which are more engaged in sensory, attention, decision making and facial recognition that were required for stimulus responses rather than for memory retention. Several studies have reported decreases in diffusion metrics or volume size in ALIC in schizophrenia (52-54), although a few did not find the same decrease (29, 52). ALIC contains fibers passing through CP, which at least in part explains the similarity in FC’s alterations between ALIC and CP (Figure 3C).

The observed associations between FC declines at WM and cognitive disturbances shown in Figure 4 are supported by other evidence. First, the positive associations with WMS-III working memory index occurred not only in cerebral bundles (e.g., UF and CGG) but also in cerebellar bundles (i.e., MCBP, ICBP, SCBP and PCT), especially in working memory II scenario. There are a few diffusion MRI studies revealing that the same working memory index or other working memory metrics correlated with fractional anisotropy at UF and CGG (25, 27), but no studies on ‘structure-working memory’ relationships at cerebellar bundles in schizophrenia. However, accumulating evidence reveals that cerebellar damage produces deficits in working memory and disturbance of prefronto-thalamo-cerebellar circuit contributes to pathophysiology of schizophrenia (55-57). Thus, we may infer that the observed association at cerebellar bundles might be a reflect of the association in cerebellum, connected via the prefronto-thalamo-cerebellar circuit. Second, the association with SCIP processing speed but not with other SCIP sub-functions (verbal learning-immediate/ delayed, working memory, verbal fluency, and global cognition) is consistent with a prior diffusion MRI study showing that degradation of WM integrity correlated with impairment of processing speed but no other functions measured by SCIP (58). In particular, the related WM bundles (i.e., SLF, UF, GCC, BCC, SCR, CGG and FXC) found in present study are also highly consistent with the findings (i.e. corpus callosum, cingulum, bundles under superior and inferior frontal gyri and precuneus) in that diffusion study (58). Third, the AX-CPT d’-context estimated the ability to discriminate target and nontarget based on context (39). Previous studies have suggested that deficits of context processing in schizophrenia associated with impaired recruitment of the dorsolateral prefrontal cortex, anterior cingulate cortex and parietal cortex (59-62). Consistently, our results revealed that the d’-context positively correlates with FC in WM bundles, such as CGG and ALIC, which are the main afferent/efferent bundles to those cortices. Finally, the fMRI task in our study is a spatial delayed-response task whose performance relies more on rehearsal (63) (equal to retention). In working memory I scenario (Figure 4B), the total response time exhibited a negative association with FCs at several WM bundles, in particular, bilateral internal capsule segments and FXC. These WM bundles mainly connect to parietal lobe, thalamus and hippocampus which were demonstrated to be activated in task of spatial working memory (64). To sum up, the above-mentioned evidence repeatedly suggests that the FC-cognition covariation in WM is likely to be a reflect of structure-cognition covariation in WM, FC-cognition covariation or structure-cognition covariation in GM that in the same circuits with WM.

Our investigation has several potential limitations. First, BOLD signals evoked by an stimuli or task in WM have been shown typically to have 3-4s latency relative to those in GM (17), and thus, in theory, the temporal correlation of WM-GM time-courses might lose some sensitivity for estimating FC values. Second, the WM atlas we used here covers the WM bundles primarily deep in the brain, some of which lack the portions extending to the surface. Matched diffusion MRI data and WM tractography will allow us to obtain more accurate WM bundles for each individual in future investigations. Third, one major WM bundle might comprise of multiple pathways connecting different cortical regions (65) and be engaged in different neural activities, so BOLD signals might be inhomogeneous over the entire WM bundle. Then averaging the time-courses over the bundle probably comprised the profile of primary neural activity.

In conclusion, FC in specific WM bundles declines in patients with schizophrenia relative to healthy controls. Alterations of FC in specific WM bundles significantly associate with impairments of neurocognition involving working memory storage/rehearsal, processing speed, and cognitive control. Both fMRI signals acquired at resting state and during a spatial working memory task are capable of revealing the FC declines and their neurocognitive associates, but the revealed results are scenario-dependent.

## Data Availability

All data are owned by authors and can be shared upon appropriate request via corresponding authors

## ACKNOWLEDMENTS AND DISCLOSURES

This work was supported by NIH grants NS093669 and NS113832 (Gore), MH102266 (Woodward), Vanderbilt Discovery Grant FF600670 (Gao), and the Charlotte and Donald Test Fund. We also thank the Advanced Computing Center for Research and Education (ACCRE) at Vanderbilt University for distributed computation.

No commercial support was received for the preparation of this manuscript. All authors report no biomedical financial interests or potential conflicts of interest.

## REFERENCES

1. Lee J, Park S (2005): Working Memory Impairments in Schizophrenia: A Meta-Analysis. Journal of Abnormal Psychology. 114: 599–611.

2. Blanchard JJ, Neale JM (1994): The neuropsychological signature of schizophrenia: Generalized or differential deficit? The American Journal of Psychiatry. 151: 40–48.

3. Heinrichs RW, Zakzanis KK (1998): Neurocognitive deficit in schizophrenia: a quantitative review of the evidence. Neuropsychology. 12: 426–445.

4. Friston KJ, Frith CD (1995): Schizophrenia: a disconnection syndrome? Clin Neurosci. 3: 89–97.

5. Andreasen NC, Nopoulos P, O’Leary DS, Miller DD, Wassink T, Flaum M (1999): Defining the phenotype of schizophrenia: cognitive dysmetria and its neural mechanisms. Biol Psychiatry. 46: 908–920.

6. Lawrie SM, Buechel C, Whalley HC, Frith CD, Friston KJ, Johnstone EC (2002): Reduced frontotemporal functional connectivity in schizophrenia associated with auditory hallucinations. Biol Psychiatry. 51: 1008–1011.

7. Kim JJ, Kwon JS, Park HJ, Youn T, Kang DH, Kim MS, et al. (2003): Functional disconnection between the prefrontal and parietal cortices during working memory processing in schizophrenia: a[15(O)]H2O PET study. Am J Psychiatry. 160: 919–923.

8. Venkataraman A, Whitford TJ, Westin CF, Golland P, Kubicki M (2012): Whole brain resting state functional connectivity abnormalities in schizophrenia. Schizophr Res. 139: 7–12.

9. Meyer-Lindenberg AS, Olsen RK, Kohn PD, Brown T, Egan MF, Weinberger DR, et al. (2005): Regionally Specific Disturbance of Dorsolateral Prefrontal–Hippocampal Functional Connectivity in Schizophrenia. Archives of General Psychiatry. 62: 379–386.

10. Andreasen NC, Paradiso S, O’Leary DS (1998): “Cognitive Dysmetria” as an Integrative Theory of Schizophrenia: A Dysfunction in Cortical-Subcortical-Cerebellar Circuitry? Schizophrenia Bulletin. 24: 203–218.

11. Woodward ND, Heckers S (2016): Mapping Thalamocortical Functional Connectivity in Chronic and Early Stages of Psychotic Disorders. Biological psychiatry. 79: 1016–1025.

12. Logothetis NK, Wandell BA (2004): Interpreting the BOLD Signal. Annual Review of Physiology. 66: 735–769.

13. Helenius J, Perkio J, Soinne L, Ostergaard L, Carano RA, Salonen O, et al. (2003): Cerebral hemodynamics in a healthy population measured by dynamic susceptibility contrast MR imaging. Acta Radiol. 44: 538–546.

14. Mazerolle EL, Gawryluk JR, Dillen KNH, Patterson SA, Feindel KW, Beyea SD, et al. (2013): Sensitivity to white matter FMRI activation increases with field strength. PloS one. 8:e58130-e58130.

15. Gore JC, Li M, Gao Y, Wu T-L, Schilling KG, Huang Y, et al. (2019): Functional MRI and resting state connectivity in white matter - a mini-review. Magnetic Resonance Imaging. 63: 1–11.

16. Wu TL, Wang F, Anderson AW, Chen LM, Ding Z, Gore JC (2016): Effects of anesthesia on resting state BOLD signals in white matter of non-human primates. Magn Reson Imaging. 34: 1235–1241.

17. Li M, Newton AT, Anderson AW, Ding Z, Gore JC (2019): Characterization of the hemodynamic response function in white matter tracts for event-related fMRI. Nature Communications. 10: 1140.

18. Gawryluk JR, Mazerolle EL, D’Arcy RCN (2014): Does functional MRI detect activation in white matter? A review of emerging evidence, issues, and future directions. Frontiers in Neuroscience. 8: 239.

19. Ding Z, Huang Y, Bailey SK, Gao Y, Cutting LE, Rogers BP, et al. (2018): Detection of synchronous brain activity in white matter tracts at rest and under functional loading. Proceedings of the National Academy of Sciences. 115: 595–600.

20. Wu T-L, Wang F, Li M, Schilling KG, Gao Y, Anderson AW, et al. (2019): Resting-state white matter-cortical connectivity in non-human primate brain. NeuroImage. 184: 45–55.

21. Gao Y, Sengupta A, Li M, Zu Z, Rogers BP, Anderson AW, et al. (2020): Functional connectivity of white matter as a biomarker of cognitive decline in Alzheimer’s disease. *medRxiv.2020.2005.2005.20091892*.

22. Gao Y, Li M, Zu Z, Rogers B, Anderson A, Ding Z, et al. (2019): Progressive degeneration of white matter functional connectivity in Alzheimer’s Disease. SPIE Medical Imaging. San Diego, California, United States.

23. Miyakawa T, Sumiyoshi S, Deshimaru M, Suzuki T, Tomonari H, Yasuoka F, et al. (1972): Electron microscopic study on schizophrenia. Acta Neuropathologica. 20: 67–77.

24. Davis KL, Stewart DG, Friedman JI, Buchsbaum M, Harvey PD, Hof PR, et al. (2003): White Matter Changes in Schizophrenia: Evidence for Myelin-Related Dysfunction. JAMA Psychiatry. 60: 443–456.

25. Nestor PG, Kubicki M, Gurrera RJ, Niznikiewicz M, Frumin M, McCarley RW, et al. (2004): Neuropsychological correlates of diffusion tensor imaging in schizophrenia. Neuropsychology. 18: 629–637.

26. Ardekani BA, Nierenberg J, Hoptman MJ, Javitt DC, Lim KO (2003): MRI study of white matter diffusion anisotropy in schizophrenia. NeuroReport. 14.

27. Kubicki M, Westin C-F, Nestor PG, Wible CG, Frumin M, Maier SE, et al. (2003): Cingulate fasciculus integrity disruption in schizophrenia: a magnetic resonance diffusion tensor imaging study. Biological Psychiatry. 54: 1171–1180.

28. Park H-J, Kubicki M, Shenton ME, Guimond A, McCarley RW, Maier SE, et al. (2003): Spatial normalization of diffusion tensor MRI using multiple channels. NeuroImage. 20: 1995–2009.

29. Paillère-Martinot ML, Caclin A, Artiges E, Poline JB, Joliot M, Mallet L, et al. (2001): Cerebral gray and white matter reductions and clinical correlates in patients with early onset schizophrenia. Schizophrenia Research. 50: 19–26.

30. Sigmundsson T, Suckling J, Maier M, Williams SCR, Bullmore ET, Greenwood KE, et al. (2001): Structural Abnormalities in Frontal, Temporal, and Limbic Regions and Interconnecting White Matter Tracts in Schizophrenic Patients With Prominent Negative Symptoms. American Journal of Psychiatry. 158: 234–243.

31. Christensen J, Holcomb J, Garver DL (2004): State-related changes in cerebral white matter may underlie psychosis exacerbation. Psychiatry Research: Neuroimaging. 130: 71–78.

32. Haijma SV, Van Haren N, Cahn W, Koolschijn PCMP, Hulshoff Pol HE, Kahn RS (2012): Brain Volumes in Schizophrenia: A Meta-Analysis in Over 18 000 Subjects. Schizophrenia Bulletin. 39: 1129–1138.

33. First MB, Gibbon M (2004): The Structured Clinical Interview for DSM-IV Axis I Disorders (SCID-I) and the Structured Clinical Interview for DSM-IV Axis II Disorders (SCID-II). Comprehensive handbook of psychological assessment, Vol 2: Personality assessment. Hoboken, NJ, US: John Wiley & Sons Inc, pp 134–143.

34. Kay SR, Fiszbein A, Opler LA (1987): The Positive and Negative Syndrome Scale (PANSS) for Schizophrenia. Schizophrenia Bulletin. 13: 261–276.

35. Wechsler D (2001): Wechsler Test Of Adult Reading. London: Pearson Education.

36. Wechsler D (1945): A Standardized Memory Scale for Clinical Use. The Journal of Psychology. 19: 87–95.

37. Purdon S (2005): The Screen For Cognitive Impairment in Psychiatry (SCIP): Administration Manual And Normative Data. Edmonton, Alberta, Canada: PNL Inc.

38. Rosvold H, Mirsky AF, Sarason I, Bransome ER, Beck LH (1956): A continuous performance test of brain damage. Journal of Consulting and Clinical Psychology. 20: 343–350.

39. Cohen JD, Barch DM, Carter C, Servan-Schreiber D (1999): Context-processing deficits in schizophrenia: converging evidence from three theoretically motivated cognitive tasks. J Abnorm Psychol. 108: 120–133.

40. Grant DA, Berg E (1948): A behavioral analysis of degree of reinforcement and ease of shifting to new responses in a Weigl-type card-sorting problem. Journal of Experimental Psychology. 38: 404–411.

41. Huang AS, Rogers BP, Anticevic A, Blackford JU, Heckers S, Woodward ND (2019): Brain function during stages of working memory in schizophrenia and psychotic bipolar disorder. Neuropsychopharmacology. 44: 2136–2142.

42. Oishi K, Faria A, Jiang H, Li X, Akhter K, Zhang J, et al. (2009): Atlas-based whole brain white matter analysis using large deformation diffeomorphic metric mapping: Application to normal elderly and Alzheimer’s disease participants. NeuroImage. 46: 486–499.

43. Lancaster JL, Woldorff MG, Parsons LM, Liotti M, Freitas CS, Rainey L, et al. (2000): Automated Talairach Atlas labels for functional brain mapping. Human Brain Mapping. 10: 120–131.

44. Power JD, Barnes KA, Snyder AZ, Schlaggar BL, Petersen SE (2012): Spurious but systematic correlations in functional connectivity MRI networks arise from subject motion. NeuroImage. 59: 2142–2154.

45. Benjamini Y, Hochberg Y (1995): Controlling the False Discovery Rate: A Practical and Powerful Approach to Multiple Testing. Journal of the Royal Statistical Society Series B (Methodological). 57: 289–300.

46. Cohen J (1988): Statistical power analysis for the behavioral sciences. 2nd ed. New York.

47. San Millán Ruíz D, Yilmaz H, Gailloud P (2009): Cerebral developmental venous anomalies: Current concepts. Annals of Neurology. 66: 271–283.

48. Lee S-H, Kubicki M, Asami T, Seidman LJ, Goldstein JM, Mesholam-Gately RI, et al. (2013): Extensive white matter abnormalities in patients with first-episode schizophrenia: A diffusion tensor imaging (DTI) study. Schizophrenia Research. 143: 231–238.

49. Skudlarski P, Schretlen DJ, Thaker GK, Stevens MC, Keshavan MS, Sweeney JA, et al. (2013): Diffusion Tensor Imaging White Matter Endophenotypes in Patients With Schizophrenia or Psychotic Bipolar Disorder and Their Relatives. American Journal of Psychiatry. 170: 886–898.

50. Gaffan D (1994): Dissociated effects of perirhinal cortex ablation, fornix transection and amygdalectomy: evidence for multiple memory systems in the primate temporal lobe. Experimental Brain Research. 99: 411–422.

51. Ennaceur A, Neave N, Aggleton JP (1997): Spontaneous object recognition and object location memory in rats: the effects of lesions in the cingulate cortices, the medial prefrontal cortex, the cingulum bundle and the fornix. Experimental Brain Research. 113: 509–519.

52. Levitt JJ, Kubicki M, Nestor PG, Ersner-Hershfield H, Westin CF, Alvarado JL, et al. (2010): A diffusion tensor imaging study of the anterior limb of the internal capsule in schizophrenia. Psychiatry research. 184: 143–150.

53. Rosenberger G, Nestor PG, Oh JS, Levitt JJ, Kindleman G, Bouix S, et al. (2012): Anterior limb of the internal capsule in schizophrenia: a diffusion tensor tractography study. Brain imaging and behavior. 6: 417–425.

54. Zhou S-Y, Suzuki M, Hagino H, Takahashi T, Kawasaki Y, Nohara S, et al. (2003): Decreased volume and increased asymmetry of the anterior limb of the internal capsule in patients with schizophrenia. Biological Psychiatry. 54: 427–436.

55. Andreasen NC, O’Leary DS, Cizadlo T, Arndt S, Rezai K, Ponto LL, et al. (1996): Schizophrenia and cognitive dysmetria: a positron-emission tomography study of dysfunctional prefrontal-thalamic-cerebellar circuitry. Proceedings of the National Academy of Sciences of the United States of America. 93: 9985–9990.

56. Wiser AK, Andreasen NC, O’Leary DS, Watkins GL, Boles Ponto LL, Hichwa RD (1998): Dysfunctional cortico-cerebellar circuits cause ‘cognitive dysmetria’ in schizophrenia. Neuroreport. 9: 1895–1899.

57. Yeganeh-Doost P, Gruber O, Falkai P, Schmitt A (2011): The role of the cerebellum in schizophrenia: from cognition to molecular pathways. Clinics (Sao Paulo, Brazil). 66 Suppl 1: 71–77.

58. Karbasforoushan H, Duffy B, Blackford JU, Woodward ND (2015): Processing speed impairment in schizophrenia is mediated by white matter integrity. Psychological medicine. 45: 109–120.

59. MacDonald Iii AW, Carter CS (2003): Event-Related fMRI Study of Context Processing in Dorsolateral Prefrontal Cortex of Patients With Schizophrenia. American Psychological Association, pp 689–697.

60. MacDonald AW, Cohen JD, Stenger VA, Carter CS (2000): Dissociating the Role of the Dorsolateral Prefrontal and Anterior Cingulate Cortex in Cognitive Control. Science. 288: 1835.

61. Lesh TA, Westphal AJ, Niendam TA, Yoon JH, Minzenberg MJ, Ragland JD, et al. (2013): Proactive and reactive cognitive control and dorsolateral prefrontal cortex dysfunction in first episode schizophrenia. NeuroImage: Clinical. 2: 590–599.

62. Barch DM, Carter CS, Braver TS, Sabb FW, MacDonald III A, Noll DC, et al. (2001): Selective deficits in prefrontal cortex function in medication-naive patients with schizophrenia. Archives of General Psychiatry 58: 280–288.

63. Leung HC, Gore JC, Goldman-Rakic PS (2002): Sustained Mnemonic Response in the Human Middle Frontal Gyrus during On-Line Storage of Spatial Memoranda. Journal of Cognitive Neuroscience. 14: 659–671.

64. van Asselen M, Kessels RP, Neggers SF, Kappelle LJ, Frijns CJ, Postma A (2006): Brain areas involved in spatial working memory. Neuropsychologia. 44: 1185–1194.

65. Schmahmann JD, Pandya DN (2006): Principles of organization of cerebral white matter tracts. Annals of Neurology. 60:S50-S50.

